# Non-invasive three-dimensional 1H-MR Spectroscopic Imaging of human brain glucose and neurotransmitter metabolism using deuterium labeling at 3T

**DOI:** 10.1101/2022.10.25.22281503

**Authors:** Fabian Niess, Lukas Hingerl, Bernhard Strasser, Petr Bednarik, Dario Goranovic, Eva Niess, Gilbert Hangel, Martin Krššák, Benjamin Spurny-Dworak, Thomas Scherer, Rupert Lanzenberger, Wolfgang Bogner

## Abstract

**Objectives:** Non-invasive, affordable, and reliable mapping of brain glucose metabolism is of critical interest for clinical research and routine application as metabolic impairment is linked to numerous pathologies e.g., cancer, dementia and depression. A novel approach to map glucose metabolism non-invasively in the human brain and separate normal oxidative from pathologic anaerobic pathways has been presented recently on experimental MR scanners using direct or indirect detection of deuterium-labeled glucose and downstream metabolites such as glutamate, glutamine and lactate.

The aim of this study was to demonstrate the feasibility to non-invasively detect deuterium labeled downstream glucose metabolites indirectly in the human brain via 3D proton (^1^H) MR spectroscopic imaging on a clinical 3T MR scanner without additional hardware.

**Materials and Methods:** This prospective, institutional review board approved study was performed in seven healthy volunteers (mean age, 31±4 years, 5 m/ 2 f) following written informed consent. After overnight fasting and oral deuterium-labeled glucose administration 3D metabolic maps were acquired every ∼4 min with ∼0.24 ml isotropic spatial resolution using real-time motion-, shim- and frequency-corrected echo-less 3D ^1^H-MR Spectroscopic Imaging. Time courses were analyzed using linear regression and non-parametric statistical tests. Deuterium labeled glucose and downstream metabolites were detected indirectly via their respective signal decrease in dynamic ^1^H MR spectra due to deuterium to proton exchange in the molecules.

**Results:** Sixty-five minutes after deuterium-labeled glucose administration, glutamate+glutamine (Glx) signal intensities decreased in gray/white matter (GM,WM) by -15±2%,(*p*=0.02)/-14±3%,(*p*=0.02), respectively. Strong negative correlation between Glx and time was observed in GM/WM (r=-0.71 *p*<0.001)/(r=-0.67,*p*<0.001) with 38±18% (*p*=0.02) steeper slopes, indicating faster metabolic activity in GM compared to WM. Other non-labeled metabolites showed no significant changes.

**Conclusion:** Our approach translates deuterium metabolic imaging to widely available clinical routine MR scanners without specialized hardware offering a safe, affordable, and versatile (other substances than glucose can be labeled) approach for non-invasive imaging of glucose and neurotransmitter metabolism in the human brain.

## Introduction

Impairment of glucose (Glc) metabolism in the human brain, i.e., a shift from aerobic Glc utilization towards anaerobic pathways (Warburg effect) has been linked to several pathologic conditions observed in e.g., ischemia and tumors(1). Anaerobic glycolysis plays an important role during early stages of dementia(2) and neuropsychiatric disorders such as schizophrenia and depression(3).

[^18^F]-Fluorodeoxyglucose ([^18^F]FDG) positron emission tomography (PET) is the current gold standard in clinical routine for assessing tissue-specific Glc uptake, but it requires invasive administration of unstable radioactive tracers and does not provide information on the dynamics of Glc downstream metabolites, e.g., oxidative neurotransmitter synthesis of glutamate (Glu) and glutamine (Gln), or anaerobic lactate production, in, e.g., tumors due to glucose trapping of [^18^F]FDG. Therefore, a fully non-invasive approach, to reliably map the brain Glc metabolism is of critical interest for clinical research and routine application (4).

Deuterium metabolic imaging (DMI) (5, 6) and quantitative exchanged label turnover (QELT) (7-10) are novel magnetic resonance techniques to non-invasively image Glc metabolism in animals and the human brain using deuterium-labeled Glc as tracer, which are able to separate healthy oxidative from pathologic anaerobic pathways, by simultaneously detecting the respective metabolic products glutamate+glutamine and lactate (4). DMI detects deuterium enrichment in the brain tissue directly via ^2^H-MRS, while, indirectly, QELT detects a decrease of signal intensities in conventional ^1^H-MR spectra due to deuterium-to-proton exchange in the respective molecule. Recently, DMI has been translated to clinical field strength of 3T(11) with a nominal isotropic resolution of 33 ml and an acquisition time of 10 min for each 3D dataset. QELT features higher SNR and spatial resolution (0.12 ml) with shorter acquisition times (∼3 min(7)) compared to DMI, while additionally detecting non-labeled metabolites, but so far has been applied in humans only at non-clinical field strength of 7T (7, 8).

Translating this QELT method to widely available clinical MR scanners (≤3T) without the need for additional expensive hardware, would offer radiologists immediate access to a fully non-invasive and affordable approach to map glucose downstream metabolism.

This study demonstrates the feasibility to non-invasively image glucose downstream metabolism in the human brain on a clinical 3T MR scanner using the QELT approach with time-resolved 3D proton (1H) MR spectroscopic imaging (MRSI) and deuterium-labeled glucose.

## Methods

### Participants

This study was approved by the ethic commission of the Medical University of Vienna and written informed consent was obtained from all participants. Seven healthy participants (mean age: 31±4years, 5 male, body mass index: 22±1kg/m^2^) were recruited consecutively at the Department of Biomedical Imaging and Image-Guided Therapy of the Medical University Vienna with the following inclusion criteria: no contraindication to 3T MRI, history of neurological or psychiatric disorders, claustrophobia, metabolic disorders, or metal implants.

### Study Protocol

The study included an MRI protocol and blood Glc sampling. MRI was performed in the morning after overnight fasting and immediately after oral tracer administration using deuterium-labeled Glc ([6,6’]-^2^H-Glc; 0.8g/kg body weight in 200ml water). Capillary puncture blood sampling from the toe (i.e., most accessible sampling site in the MR scanner) was performed six times over the course of ∼90 min using a standard strip glucometer.

### MRI Protocol

All measurements were conducted at a clinical routine 3T MR system (Prisma-FIT) using a 64-receive head coil (Siemens Healthineers, Erlangen). Preparation scans included an automated alignment localizer followed by echo planar imaging reference scans to set up the volumetric navigator sequence used for real-time motion correction(12). A previously developed 3D echo-less (FID) MRSI sequence with automatic interleaved real-time motion-, shim- and frequency drift-correction and fast concentric ring trajectory readout (13) was used to acquire 14 consecutive 3D datasets over the course of ∼60min: 0.8ms acquisition delay, 950ms repetition time, 0.24ml isotropic nominal voxel volume (matrix size: 32×32×21, field of view: 200×200×130mm^3^, volume of interest: 200×200×55 mm^3^ centered around the posterior cingulate region) and 4:14min acquisition time per repetition (for details see table, Supplemental Digital Content 1 (14)).

Following the MRSI scan a 3D T1-weighted magnetization-prepared rapid gradient echo readout (MPRAGE) scan was performed: 1800ms repetition time, 2.27ms echo time, 900ms inversion time, 1mm^3^ isotropic nominal voxel volume, 3-fold parallel acquisition acceleration (GRAPPA), and 2:38min acquisition time.

### Data Reconstruction

An in-house developed software pipeline (MATLAB R2021, Python3.10) was used for automatic data processing including: k-space in-plane convolutional gridding(15), noise-decorrelation, channel-wise lipid decontamination(16), coil combination(17) and spectral fitting (4.2-1.8 ppm) using LCModel (version 6.3). Quantification results of voxels which did not fulfil the quality control criteria, i.e., full-width-half-maximum <0.1ppm, signal-to-noise ratio >15, Cramer-Rao Lower Bounds <20% were excluded from the analysis.

### Metabolite Quantification

A modified basis set was used featuring 17 typical metabolites (i.e., creatine, glycerophosphocholine, glutathione, myo-inositol, N-acetylaspartate, N-Acetylaspartylglutamate, phosphocholine, phosphocreatine, taurine, aspartate, gamma-aminobutyric acid, Glc≤, Glc≤glutamate, glutamine, lactate) of neurochemical profile (including measured macromolecular contributions)(18), but this work focuses on few selected metabolites of interest: Glutamate+Glutamine (Glu+Gln: Glx), total creatine (tCr) and total N-acetylaspartate (tNAA). During metabolic utilization of deuterium-labeled Glc (([6,6’]-^2^H-Glc), downstream metabolites incorporate deuterium at specific carbon positions. In case of oxidatively synthesized Glu and Gln deuterium labeling occurs only at the 4^th^ carbon position (Glu_4_: 2.34ppm, Gln_4_: 2.44ppm), eventually leading to a signal intensity decrease of the respective resonances, while resonances associated with protons at the 2^nd^ and 3^rd^ position (Glu_2_: 3.75ppm, Glu_3_: 2.10ppm, Gln_2_: 3.77ppm and Gln_3_: 2.13ppm) remain stable.

Therefore, resonances representing Glu and Gln in the basis set were separated into Glu_4_, Glu_2+3_ and Gln_4_, Gln_2+3_ and then summed to Glx_4_ and Glx_2+3_, representing labeled and unlabeled resonances, respectively. Separation was performed by simulating the fully deuterated state of Glu and Gln (equals Glu_2+3_ and Gln_2+3_: both protons at the 4^th^ carbon position are replaced with deuterons) and subtracting it from regular Glu and Gln to obtain Glu_4_ and Gln_4_. Therefore, a linear combination equals regular Glu and Gln molecules taking J-coupling effects into account. Similarly, only the 6^th^ carbon position of Glc is labeled and, therefore, Glc_6_ (labeled) and Glc_1-5_ (unlabeled) components were introduced. Labeling of Glc on other carbon positions than [6,6] are lost during metabolic utilization and are not detectable. Metabolite concentration estimates are given in arbitrary units (a.u.) throughout the whole manuscript.

### Time course analysis

3D metabolic maps were created for all 14 time points representing the metabolic dynamics over ∼60min with a high time resolution of ∼4min. Maps were co-registered to anatomical T1-weighted images. Regional segmentation for gray and white matter(GM and WM), was automatically performed using the FAST algorithm(19) on T1-weighted 3D images and down-sampled to 32×32 using MINC tools (MINC tools, v2.0, McConnell Brain Imaging Center, Montreal, QC, Canada). Partial volume effects were minimized using a threshold of 80% for GM and WM voxels, respectively.

Temporal metabolite signal evolution was investigated voxel-wise and over the whole GM or WM in each participant given as mean±standard deviation. To estimate temporal stability of metabolite concentration fits, coefficients of variation were calculated over the entire MRSI scan (60 min and 14 time points) for stable non-labeled metabolites (Glx_2+3_, tCr, tNAA).

### Statistical Analysis

Linear regression analysis was performed between time and quantified metabolite signals, e.g., deuterium-labeled resonances (Glx_4_). Differences between the first and last time points and between GM and WM groups were evaluated using Wilcoxon signed-rank test and Mann-Whitney U test between segmented GM and WM voxels. The statistical significance threshold was *p*<0.05. Linear fitting and statistical tests were performed using Python3.10 (www.python.org, packages: scipy.stats).

## Results

### Study Protocol

All preparation scans were completed 6±2min after tracer administration. The subsequent MRSI sequence acquired 3D datasets consecutively every ∼4 minutes without interruption or operator interaction. Capillary blood sampling and analysis was performed in five participants.

### Time course analysis

A strong negative correlation with time was observed for Glx_4_ in both GM (r=-0.71,*p*<0.001) and WM (r=-0.67,*p*<0.001) with 38±18% steeper slopes in GM compared to WM (*p*=0.02) representing faster signal decay (Figure 1a). Individual results of linear regression analysis are listed in Table 1.

**Table 1:** Linear regression results per participant and region.

| Metabolite | slopes / [a.u.]*: | correlation coeff.* [r] | p value* |
| --- | --- | --- | --- |
| Glx<sub>4</sub> |  |  |  |
| Participant 1 | -41.8/-39.4 | -0.96/-0.97 | <0.001/<0.001 |
| Participant 2 | -46.2/-30.0 | -0.83/-0.74 | <0.001/0.002 |
| Participant 3 | -59.6/-50.5 | -0.94/-0.93 | <0.001/<0.001 |
| Participant 4 | -42.2/31.2 | -0.93/-0.90 | <0.001/<0.001 |
| Participant 5 | -53.4/-38.6 | -0.94/-0.95 | <0.001/<0.001 |
| Participant 6 | -55.1/ -37.2 | -0.94/-0.95 | <0.001/<0.001 |
| Participant 7 | -46.2/-34.2 | -0.95/-0.93 | <0.001/<0.001 |

Note.—GM = gray matter, WM = white matter, Glx = glutamate+glutamine
\* Data are from gray and white matter (GM/WM)

**Figure 1:**
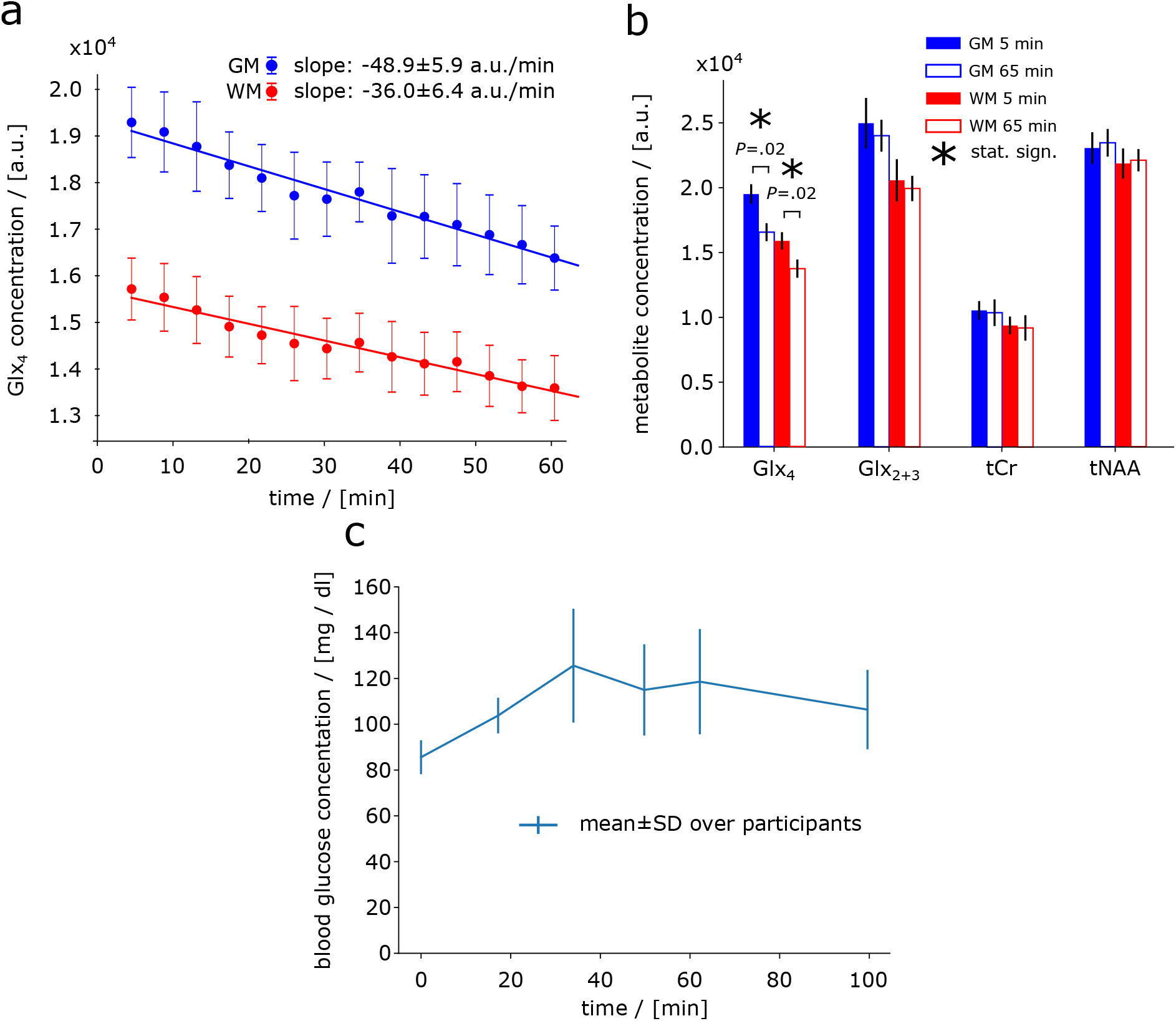
Time courses of deuterium labeled glutamate+glutamine (Glx_4_) signal intensities (from whole gray and white matter averaged over all participants. A decrease of Glx_4_ over time can be observed in both gray and white matter (GM, WM). Signal intensity decrease of Glx_4_ was 38±18% faster in GM compared to WM. First and last measurement (first at ∼5. min, last at ∼65. min) of deuterium labeled (Glx_4_) and unlabeled (Glx_2+3_, tCr, tNAA) resonances shown as regional means over whole GM and WM and over all participants (b). Averaged time course of blood Glc concentration over all participants increasing from 86±7 mg/dl to a maximum of 126±25 mg/dl at 34 min after tracer administration before gradually decreasing towards baseline (c).

Sixty-five minutes after tracer administration, Glx_4_ had decreased by -15±2% (*p*=0.02) and -13±3%, (*p*=0.02) compared to the initial time point in GM and WM, respectively, while no changes were found for Glx_2+3_, tCr, or tNAA (GM/WM: Glx_2+3_ *p*=0.08/0.16, tCr *p*=0.47/0.30, tNAA: *p*=0.16/0.81) (Figure 1b). Blood Glc concentration increased from 86±7mg/dl at baseline to a maximum of 126±25mg/dl (*p*=0.068) at 34 min after tracer administration followed by a gradual decrease towards baseline (Figure 1c).

Non-labeled metabolite resonances, e.g., Glx_2+3_, tCr and tNAA featured high temporal stability with coefficients of variation <2% (Glx_2+3_:1.9±0.6%, tCr:1.5±0.8% and tNAA:1.1±1%).

Representative axial Glx_4_ maps are shown for all time points from one representative subject (Figure 2). A continuous signal intensity decrease over time due to deuterium labeling was visually discernable. Metabolic map time courses from all participants are shown in figure, Supplemental Digital Content 1.

**Figure 2:**
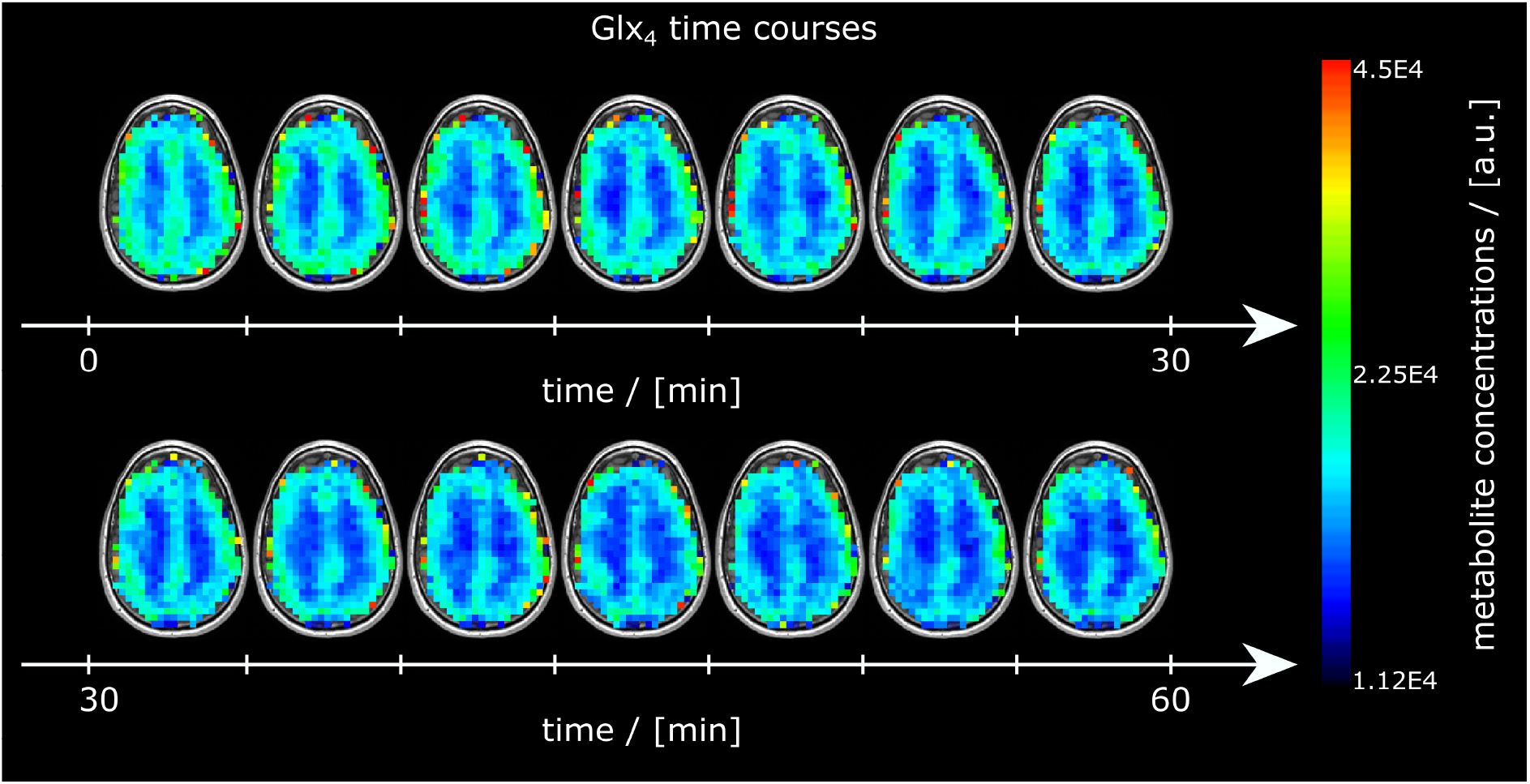
Time courses of axial glutamate+glutamine (Glx_4_) maps from one representative subject over the entire measurement visualizing the image intensity decrease over time due to deuterium labeling.

Voxel-wise linear fitting yielded moderate to strong (r< -0.5, *p*<0.05) negative correlation between Glx_4_ signal intensity and time in 50% of GM and 44% of WM voxels from a 3D dataset of one representative participant (Figure 3). Slopes of the linear regression show a visible GM/WM contrast with 32% steeper slopes for Glx_4_ (*p*<0.001) between GM and WM representing a faster signal intensity decay, i.e., higher metabolic activity.

**Figure 3:**
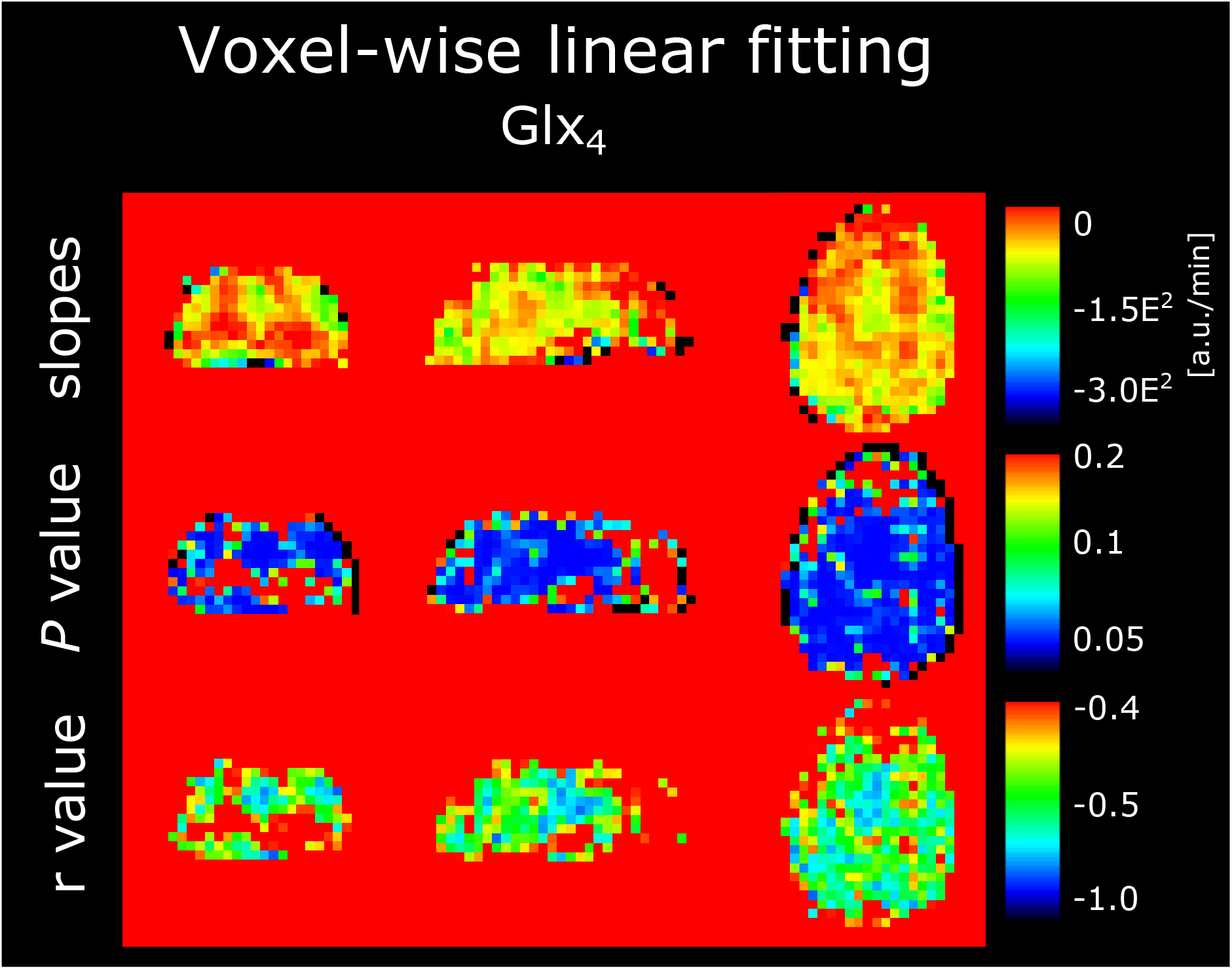
Voxel-wise linear fitting between time and deuterium labeled metabolites, i.e., glutamate+glutamine (Glx_4_) shown from one representative participant featuring slopes, *P* values r values (top,middle, bottom row respectively) for each voxel separately of the whole 3D volume. The slope map shows a contrast between GM and WM with 32% steeper slopes for Glx_4_ (p<0.001) in GM versus WM. Linear fitting was statistically significant (*p* < 0.05) with correlation (r < -0.5) in 50% and 44% of GM and WM voxels, respectively.

For illustration purposes subtraction spectra (first–last measurement) of averaged GM and WM voxels of one representative subject were calculated to visualize the signal drop of the Glx_4_ resonance 65 min after tracer administration as an increase of deuterium-labeled resonances only, similar to DMI (Figure 4). Subtraction spectra showed Glx_4_ and Glc_6_ resonances. However, spectral fitting was performed voxel wise and without spectral subtraction.

**Figure 4:**
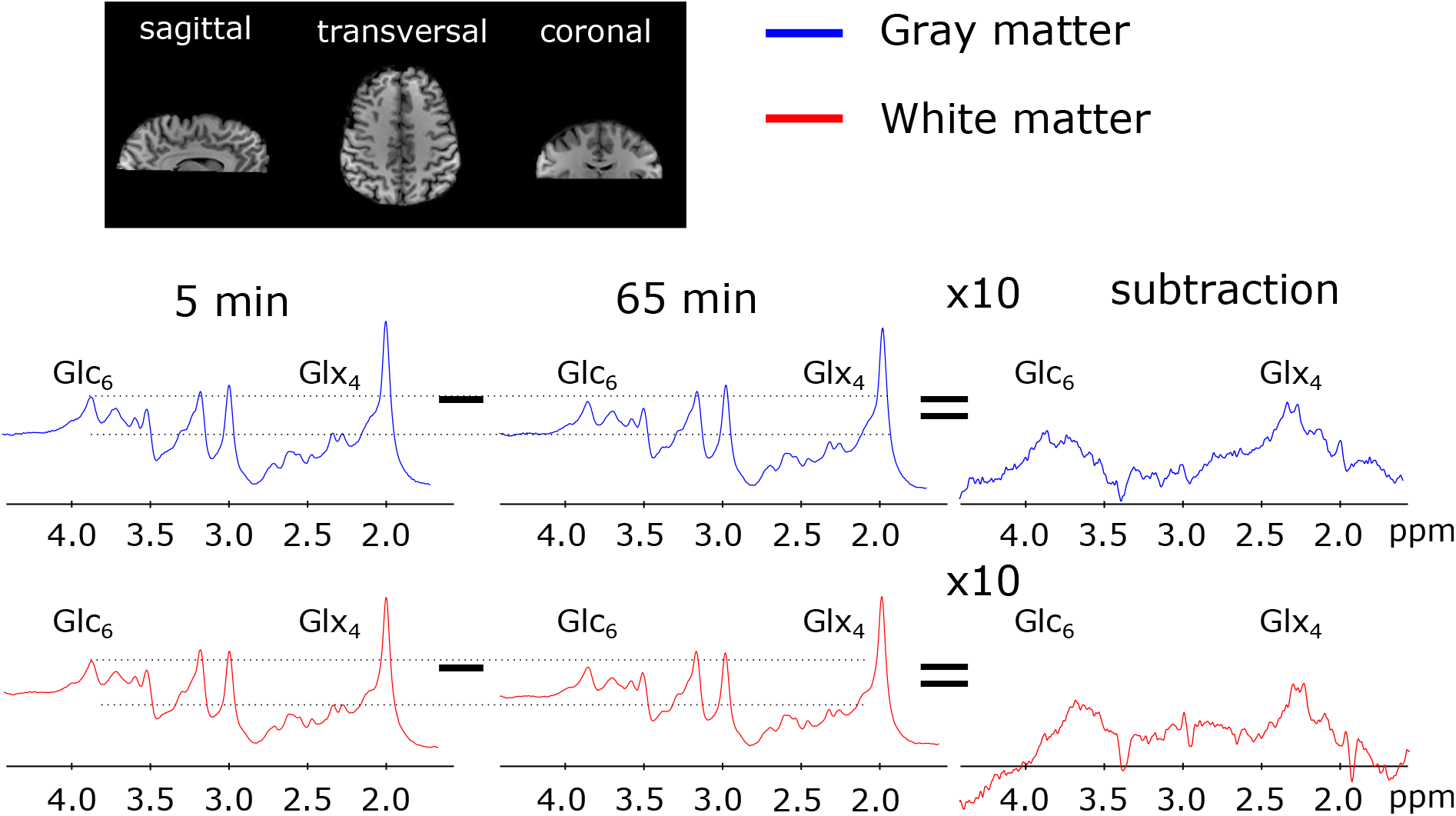
Averaged spectra over selected voxels in gray and white matter 5 min and 65 min after deuterium-labeled Glc administration visualizing a decrease in the Glx_4_ and Glc_6_ resonance, while other resonances were stable. Corresponding subtraction spectra showed an increase of the Glx_4_ and Glc_6_ resonance.

More than 90% of GM and WM voxels fulfilled our standardized quality criteria. Sample spectra of single GM and WM voxels from the first and last time points of one representative subject are shown in figure, Supplemental Digital Content 2, with their respective metabolite concentrations and CRLBs see table, Supplemental Digital Content 2.

## Discussion

In this study we have targeted the need for a fully non-invasive clinically applicable technique to map the dynamics of glucose metabolism in the human brain. We demonstrated the feasibility of our MR technique to image oxidative downstream glucose metabolism almost over the entire human cerebrum, using orally administered deuterium labeled glucose on a clinical routine 3T MR system. The ultra-short echo time of the 3D FID-MRSI method minimizes J-evolution for metabolites such as Glx and improves the SNR compared to spin-echo approaches. High SNR and integrated real-time motion correction (13) provided a high temporal stability reflected by a coefficient of variation of under two percent over the course of 60 min (14 time points) for regional averages of non-labeled metabolites, i.e., Glx_2+3_, tCr and tNAA. This allowed for a reliable detection of 10-20% changes in signal amplitude for labeled metabolites (Glx_4_) due to deuterium enrichment even for voxel-wise analysis. However, regional averaging over multiple voxels improved the robustness of the results.

A proof, that the decrease of Glx_4_ is not an artifact or systematic error of the method and indeed reflects physiological incorporation of deuterium into downstream metabolites, has been performed in a previous study by assessing the test-retest repeatability and control measurements using regular glucose (dextrose), employing a similar MR sequence (without motion correction) at 7T (7). Additionally, results were compared to DMI data acquired from the same cohort of subjects and on the same MR scanner. Therefore, no test-retest repeatability, reproducibility and control measurements were performed in this study. The enrichment of labeled Glx_4_ in our study is in good agreement with literature (6, 8-10, 20).

A thirty-three percent faster metabolic activity in GM compared to WM in our study is in line with [^18^F]FDG-PET literature values, which reported ∼33 % higher oxidative Glc consumption in GM than in WM (21-23), but lower than differences reported in the rate of TCA cycle(68%) measured in ^13^C-studies(24, 25). A visible decrease of the Glc_6_ resonance could be observed in Figure 4, but reliable fitting of the Glc resonances (labeled Glc_6_ and unlabeled Glc_1-5_) was not possible on voxel-wise level with comparable quality, presumably due to overlapping resonances in the region 3.5-4.0 ppm. For illustration purposes metabolic map time courses without quality control thresholds are shown for all subjects in figure, Supplemental Digital Content 3. To calculate quantitative flux parameters of glucose metabolism, detection of the glucose uptake by detecting the decrease of Glc_6_ resonances needs to be improved but the main aim of this study was a qualitative assessment of oxidative downstream metabolites, which directly reflects glucose utilization.

In contrast to a direct detection of labeled substrates using DMI (^2^H-MRS/MRSI), ^13^C-MRS, or [^18^F]FDG-PET the indirect ^1^H-based QELT approach allows for simultaneous detection of labeled and unlabeled metabolites to quantify an extended neurochemical profile and requires no additional hardware or radioactive tracers (8-10). In comparison to recent publications presenting QELT MRSI results in animals (9) and humans (7, 8, 10) at ultra-high field(≥7T) research systems, our approach provides motion-corrected 3D metabolite maps with high temporal stability (26), which were acquired with high spatial and temporal resolution on widely available clinical 3T MR scanners. Further improvements in spatial and temporal stability can be anticipated considering the wide range of state-of-the-art undersampling or super-resolution techniques in MRSI (27-29).

Similar to DMI, a separation of healthy oxidative and pathologic anaerobic metabolic pathways, via simultaneous detection of Glx and lactate is theoretically feasible using ^1^H-MRSI. However, only 2% of glucose is anaerobically converted in the healthy human brain and, thus, we could not reliably quantify lactate concentrations in our study. Future applications could focus on pathologies involving anaerobic glucose utilization, e.g., types of brain tumors, but as this introduces additional technical challenges, this would extend the scope of this study. A residual contamination from subcutaneous fat outside of the brain due to e.g., a suboptimal point spread function(15) additionally limits the detection of lactate. Fat suppression (30, 31), volume selective excitation using spin-echo approaches or higher spatial resolution (32), could improve lactate detection in future studies involving pathologies with high lactate production (33).

While our study shows only results for investigating glucose metabolism, deuterium-labeling is not limited to glucose. A range of metabolites can be deuterium-labeled, e.g., choline, acetate (34) to study different metabolic pathways and pathologies—some of them even simultaneously using multiple tracers, e.g., in tumors (35). This adds additional flexibility for designing clinical research studies. Deuterium labeled glucose ([6,6’]-^2^H-Glc) is a safe and stable tracer and the price of the administered dose was under 1000 $ for each subject. However, this price is expected to drop upon mass production and the approval of cheaper approaches to synthesize e.g., deuterated glucose (36, 37).

Oral administration was preferred over intravenous injection in favor of participant comfort, and expected higher isotopic enrichment of Glx_4_ due to slower absorption and release of deuterium-labeled Glc as presented in ^13^C studies (20). Hence, a delayed response after oral Glc uptake, a non-steady-state of blood Glc and relatively short measurement time limits possible calculation of quantitative flux rates (38), which was not the aim of this study. Intravenous tracer injection would allow additional blood sampling to estimate isotopic enrichment time courses as an input function to overcome those challenges in future studies (20). A previous study using DMI showed that the increase of deuterium labeled Glx features an approximately linear behavior in the first 60 min(7), therefore, our study used linear fitting as an approximation to estimate the dynamics of Glx, but this approach does not reflect real quantitative turnover rates. Other models, such as exponential fitting has been applied previously, but require longer scan times (e.g., 2-3 hours) and the assessment of a true baseline. However, whether accurately quantifying flux rates via a lengthy acquisition provides sufficient added diagnostic value in patients remains to be shown.

This study demonstrates not only the feasibility of dynamic three-dimensional ^1^H-MRSI to non-invasively image glucose downstream metabolism in the human brain using deuterium-labeled glucose at widely available clinical 3T MR scanners, but suggests the possibility to map several other deuterated downstream metabolites of clinical interest.

## Data Availability

All data produced in the present study are available upon reasonable request to the authors

## Supplemental Digital Content

### Niess_SDC.pdf

Supplemental Digital Content Table 1

Supplemental Digital Content Table 2

Supplemental Digital Content Figure 1

Supplemental Digital Content Figure 2

Supplemental Digital Content Figure 3

## References

1. Koppenol WH, Bounds PL, Dang CV. Otto Warburg’s contributions to current concepts of cancer metabolism. Nat Rev Cancer. 2011;11(5):325–37.

2. Norat P, Soldozy S, Sokolowski JD, et al. Mitochondrial dysfunction in neurological disorders: Exploring mitochondrial transplantation. NPJ Regen Med. 2020;5(1):22.

3. Manji H, Kato T, Di Prospero NA, et al. Impaired mitochondrial function in psychiatric disorders. Nat Rev Neurosci. 2012;13(5):293–307.

4. Kumar M, Nanga RPR, Verma G, et al. Emerging MR Imaging and Spectroscopic Methods to Study Brain Tumor Metabolism. Front Neurol. 2022;13:789355.

5. De Feyter HM, Behar KL, Corbin ZA, et al. Deuterium metabolic imaging (DMI) for MRI-based 3D mapping of metabolism in vivo. Sci Adv. 2018;4(8):eaat7314.

6. Ruhm L, Avdievich N, Ziegs T, et al. Deuterium metabolic imaging in the human brain at 9.4 Tesla with high spatial and temporal resolution. Neuroimage. 2021;244:118639.

7. Bednarik P, Goranovic D, Svatkova A, et al. Deuterium labeling enables non-invasive 3D proton MR imaging of glucose and neurotransmitter metabolism in the human brain at 7T. Nat Biomed Eng. 2022.

8. Cember ATJ, Wilson NE, Rich LJ, et al. Integrating (1)H MRS and deuterium labeled glucose for mapping the dynamics of neural metabolism in humans. Neuroimage. 2022;251:118977.

9. Rich LJ, Bagga P, Wilson NE, et al. (1)H magnetic resonance spectroscopy of (2)H-to-(1)H exchange quantifies the dynamics of cellular metabolism in vivo. Nat Biomed Eng. 2020;4(3):335–42.

10. Ruhm L, Ziegs T, Wright AM, et al. Dynamic observation of <sup>2</sup>H labeled compounds in the human brain with <sup>1</sup>H versus <sup>2</sup>H magnetic resonance spectroscopy at 9.4T. bioRxiv. 2022:2022.01.24.477582.

11. Kaggie JD, Khan AS, Matys T, et al. Deuterium metabolic imaging and hyperpolarized (13)C-MRI of the normal human brain at clinical field strength reveals differential cerebral metabolism. Neuroimage. 2022;257:119284.

12. Bogner W, Hess AT, Gagoski B, et al. Real-time motion- and B0-correction for LASER-localized spiral-accelerated 3D-MRSI of the brain at 3T. Neuroimage. 2014;88:22–31.

13. Moser P, Eckstein K, Hingerl L, et al. Intra-session and inter-subject variability of 3D-FID-MRSI using single-echo volumetric EPI navigators at 3T. Magn Reson Med. 2020;83(6):1920–9.

14. Lin A, Andronesi O, Bogner W, et al. Minimum Reporting Standards for in vivo Magnetic Resonance Spectroscopy (MRSinMRS): Experts’ consensus recommendations. NMR Biomed. 2021;34(5):e4484.

15. Hingerl L, Bogner W, Moser P, et al. Density-weighted concentric circle trajectories for high resolution brain magnetic resonance spectroscopic imaging at 7T. Magn Reson Med. 2018;79(6):2874–85.

16. Bilgic B, Chatnuntawech I, Fan AP, et al. Fast image reconstruction with L2-regularization. J Magn Reson Imaging. 2014;40(1):181–91.

17. Strasser B, Chmelik M, Robinson SD, et al. Coil combination of multichannel MRSI data at 7 T: MUSICAL. NMR Biomed. 2013;26(12):1796–805.

18. Povazan M, Hangel G, Strasser B, et al. Mapping of brain macromolecules and their use for spectral processing of (1)H-MRSI data with an ultra-short acquisition delay at 7 T. Neuroimage. 2015;121:126–35.

19. Jenkinson M, Beckmann CF, Behrens TE, et al. Fsl. Neuroimage. 2012;62(2):782–90.

20. Moreno A, Bluml S, Hwang JH, Ross BD. Alternative 1-(13)C glucose infusion protocols for clinical (13)C MRS examinations of the brain. Magn Reson Med. 2001;46(1):39–48.

21. Hyder F, Fulbright RK, Shulman RG, Rothman DL. Glutamatergic function in the resting awake human brain is supported by uniformly high oxidative energy. J Cereb Blood Flow Metab. 2013;33(3):339–47.

22. Hyder F, Rothman DL. Quantitative fMRI and oxidative neuroenergetics. Neuroimage. 2012;62(2):985–94.

23. Yu Y, Herman P, Rothman DL, et al. Evaluating the gray and white matter energy budgets of human brain function. J Cereb Blood Flow Metab. 2018;38(8):1339–53.

24. Pan JW, Stein DT, Telang F, et al. Spectroscopic imaging of glutamate C4 turnover in human brain. Magn Reson Med. 2000;44(5):673–9.

25. Shulman RG, Rothman DL, Behar KL, Hyder F. Energetic basis of brain activity: implications for neuroimaging. Trends Neurosci. 2004;27(8):489–95.

26. Andronesi OC, Bhattacharyya PK, Bogner W, et al. Motion correction methods for MRS: experts’ consensus recommendations. NMR Biomed. 2021;34(5):e4364.

27. Bogner W, Otazo R, Henning A. Accelerated MR spectroscopic imaging-a review of current and emerging techniques. NMR Biomed. 2021;34(5):e4314.

28. Iqbal Z, Nguyen D, Hangel G, et al. Super-Resolution (1)H Magnetic Resonance Spectroscopic Imaging Utilizing Deep Learning. Front Oncol. 2019;9:1010.

29. Hangel G, Jain S, Springer E, et al. High-resolution metabolic mapping of gliomas via patch-based super-resolution magnetic resonance spectroscopic imaging at 7T. Neuroimage. 2019;191:587–95.

30. Tkac I, Deelchand D, Dreher W, et al. Water and lipid suppression techniques for advanced (1) H MRS and MRSI of the human brain: Experts’ consensus recommendations. NMR Biomed. 2021;34(5):e4459.

31. Hangel G, Strasser B, Povazan M, et al. Lipid suppression via double inversion recovery with symmetric frequency sweep for robust 2D-GRAPPA-accelerated MRSI of the brain at 7 T. NMR Biomed. 2015;28(11):1413–25.

32. Hangel G, Strasser B, Povazan M, et al. Ultra-high resolution brain metabolite mapping at 7 T by short-TR Hadamard-encoded FID-MRSI. Neuroimage. 2018;168:199–210.

33. Maudsley AA, Andronesi OC, Barker PB, et al. Advanced magnetic resonance spectroscopic neuroimaging: Experts’ consensus recommendations. NMR Biomed. 2021;34(5):e4309.

34. Polvoy I, Qin H, Flavell RR, et al. Deuterium Metabolic Imaging-Rediscovery of a Spectroscopic Tool. Metabolites. 2021;11(9).

35. Veltien A, van Asten J, Ravichandran N, et al. Simultaneous Recording of the Uptake and Conversion of Glucose and Choline in Tumors by Deuterium Metabolic Imaging. Cancers (Basel). 2021;13(16).

36. Fujiwara Y, Iwata H, Sawama Y, et al. Method for regio-, chemo- and stereoselective deuterium labeling of sugars based on ruthenium-catalyzed C-H bond activation. Chem Commun (Camb). 2010;46(27):4977–9.

37. Li W, Rabeah J, Bourriquen F, et al. Scalable and selective deuteration of (hetero)arenes. Nat Chem. 2022;14(3):334–41.

38. Kreis F, Wright AJ, Hesse F, et al. Measuring Tumor Glycolytic Flux in Vivo by Using Fast Deuterium MRI. Radiology. 2020;294(2):289–96.

