## Supplemental Tables 1-2 and Supplemental Figures 1-3 for "Non-invasive three-dimensional 1H-MR Spectroscopic Imaging of human brain glucose and neurotransmitter metabolism using deuterium labeling at 3T"

| Minimum Reporting Standards in MR Spectroscopy checklist (according to Lin et al. NMR Biomed 2021) | |
|---|---|
| **1. Hardware** | |
| **a. Field strength [T]** | 3 |
| **b. Manufacturer** | Siemens |
| **c. Model (software version if available)** | Prisma Fit |
| **d. RF coils: nuclei (transmit/ receive), number of channels, type, body part** | 1H TX/RX 64 channels, head, Siemens |
| **e. Additional hardware** | N/A |
| **2. Acquisition** | |
| **a. Pulse sequence** | FID-CRT MR spectroscopic imaging |
| **b. Volume of Interest (VOI) locations** | The excited 55 mm-thick slab was centered around the posterior cingulate region. |
| **c. Nominal VOI size [$cm^3$, $mm^3$]** | 200×200×55 $mm^3$ |
| **d. Repetition Time (TR), Echo Time (TE) [ms, s]** | TR=950 ms / 0.8 ms acquisition delay |
| **e. Total number of Excitations or acquisitions per spectrum** | 1 average |
| **In time series for kinetic studies** | N/A |
| **i. Number of Averaged spectra (NA) per time-point** | N/A |
| **ii. Averaging method (e.g. block-wise or moving average)** | N/A |
| **iii. Total number of spectra (acquired / in time-series)** | N/A |
| **f. Additional sequence parameters (spectral width in Hz, number of spectral points, frequency offsets); If STEAM: Mixing Time TM; If MRSI: 2D or 3D, FOV in all directions, matrix size, acceleration factors** | Bandwidth: 1325 Hz, 588 spectral points, MRSI: 3D, FOV 200×200x130 $mm^2$, |
| **g. Water Suppression Method** | WET |
| **h. Shimming Method, reference peak, and thresholds for "acceptance of shim" chosen** | Standard shim + manual adjustment, water peak < 30 Hz |
| **i. Triggering or motion correction method (respiratory, peripheral, cardiac triggering, incl. device used and delays)** | N/A |
| **3. Data analysis methods and outputs** | |
| **a. Analysis software** | LCModel 6.3-1 |
| **b. Processing steps deviating from quoted reference or product** | N/A |
| **c. Output measure (e.g. absolute concentration, institutional units, ratio)** | institutional units, ratio |
| **d. Quantification references and assumptions, fitting model assumptions** | Simulated in NMRScope-B, macromolecular background |
| **4. Data Quality** | |
| **a. Reported variables (SNR, Linewidth (with reference peaks))** | SNR was calculated using the pseudoreplica method, and linewidth as FWHM of the NAA fit |
| **b. Data exclusion criteria** | CRLBs >20% for tNAA, tCr and Glu+Gln (Glx) |
| **c. Quality measures of postprocessing Model fitting (e.g. CRLB, goodness of fit, SD of residual)** | CRLB |
| **d. Sample Spectrum** | See Supplemental Digital Content Figure 2 |

**Supplemental Digital Content Table 1:** Minimum Reporting Standards for in vivo MR Spectroscopy

Note. – CRLB = Cramér-Rao lower bounds; FID = free induction decay; FOV = field of view; FWHM = full-width-at-half-maximum; Glu = Glutamate; Gln = Glutamine; tNAA = total N-acetylaspartate; SNR = signal-to-noise ratio; tCr = total creatine; VOI = volume of interest.

| | Gray matter voxel concentration [a.u.] (CRLB [%]) | | White matter voxel concentration [a.u.] (CRLB [%]) | |
|---|---|---|---|---|
| Metabolite | 5 min | 65 min | 5 min | 65 min |
| tCr | 1.07E4 (4%) | 1.00E4 (5%) | 7.47E3 (5%) | 6.75E3 (4%) |
| tNAA | 2.25E4 (3%) | 2.25E4 (3%) | 1.90E4 (3%) | 2.03E4 (2%) |
| Glx4* | 2.10E4 (7%) | 1.53E4 (10%) | 1.09E4 (10%) | 9.00E3 (11%) |
| Glx23 | 2.68E4 (5%) | 2.76E4 (5%) | 1.36E4 (8%) | 1.44E4 (6%) |
| Glu4* | 1.44E4 (7%) | 1.06E4 (10%) | 7.77E3 (10%) | 6.21E3 (12%) |
| Glu23 | 1.83E4 (7%) | 1.82E4 (8%) | 8.51E3 (11%) | 6.60E3 (16%) |
| Gln4* | 6.55E3 (12%) | 4.63E3 (17%) | 3.11E3 (18%) | 2.80E3 (19%) |
| Gln23 | 8.46E3 (13%) | 9.36E3 (13%) | 5.11E3 (17%) | 7.76E3 (12%) |
| Cr | 7.13E3 (13%) | 5.84E3 (17%) | 5.58E3 (12%) | 4.00E3 (15%) |
| PCr | 3.60E3 (24%) | 4.19E3 (24%) | 1.89E3 (34%) | 2.75E3 (22%) |
| NAA | 2.12E4 (3%) | 2.21E4 (3%) | 1.69E4 (3%) | 1.68E4 (3%) |
| NAAG | 1.30E3 (30%) | 4.15E2(111%) | 2.17E3 (14%) | 3.54E3 (9%) |

*deuterium labeled
unlabeled

**Supplemental Digital Content Table 2:** LCModel spectral fit output of a single representative GM and WM voxel for selected metabolites from the first (5 min) and last (60 min) time point after D-glucose ingestion, respectively. Respective spectra and LCModel fits are shown in Supplemental Digital Content Figure 2.

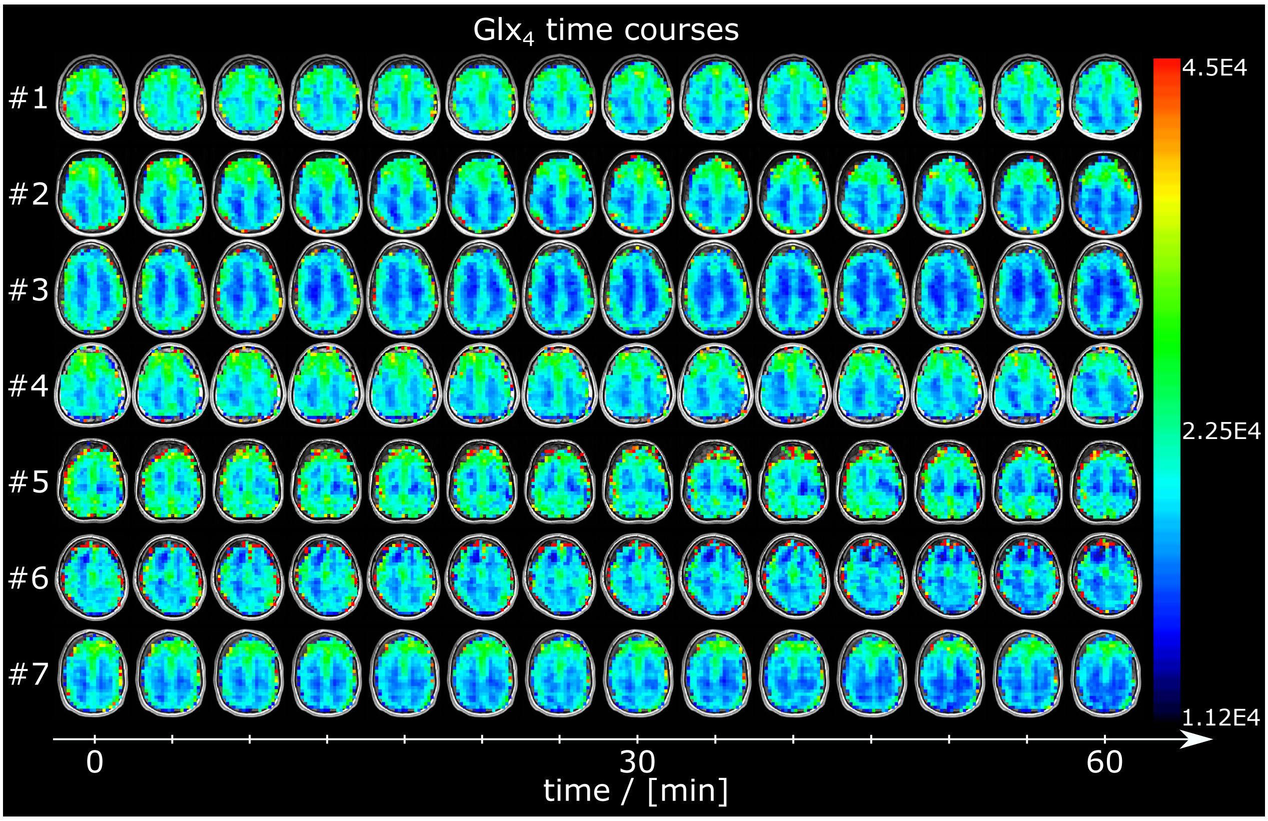


**Supplemental Digital Content Figure 1:** Time courses of axial glutamate+glutamine ($Glx_4$) maps from all subjects over the entire measurement visualizing the image intensity decrease over time due to deuterium labeling.

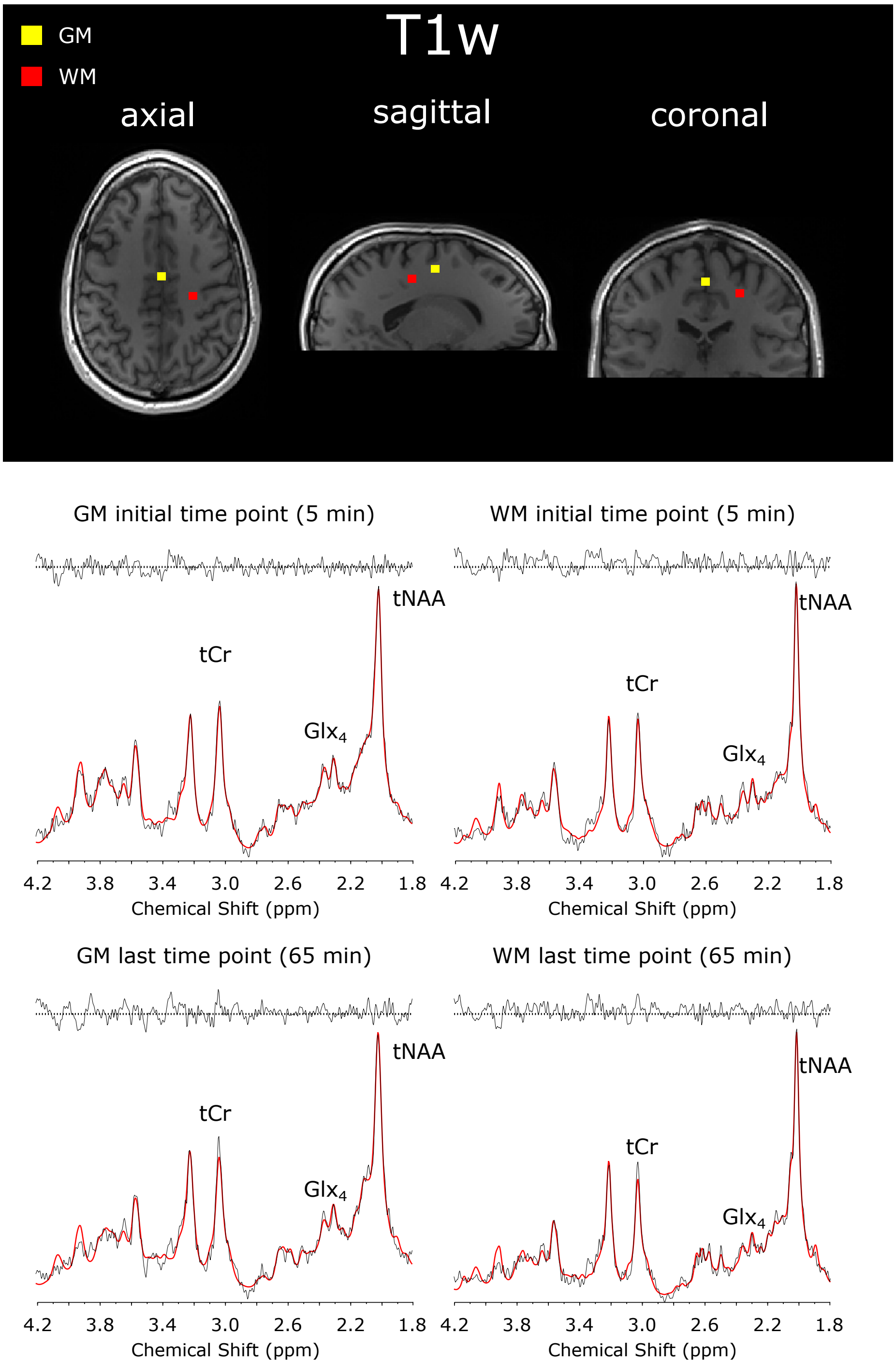


**Supplemental Digital Content Figure 2:** Sample spectra (black) and LCModel fit (red) from a single gray matter (GM) and white matter (WM) voxel of one representative volunteer at the beginning (5 min) and the end (65 min) of the MRSI scan. Spectra were first-order corrected for illustration purposes. Relevant metabolites are total creatine (tCr), glutamate+glutamine (Glx) and total N-acetylaspartate (tNAA). Spectral concentration fits of these particular voxels are shown in Supplemental Digital Content Table 1.

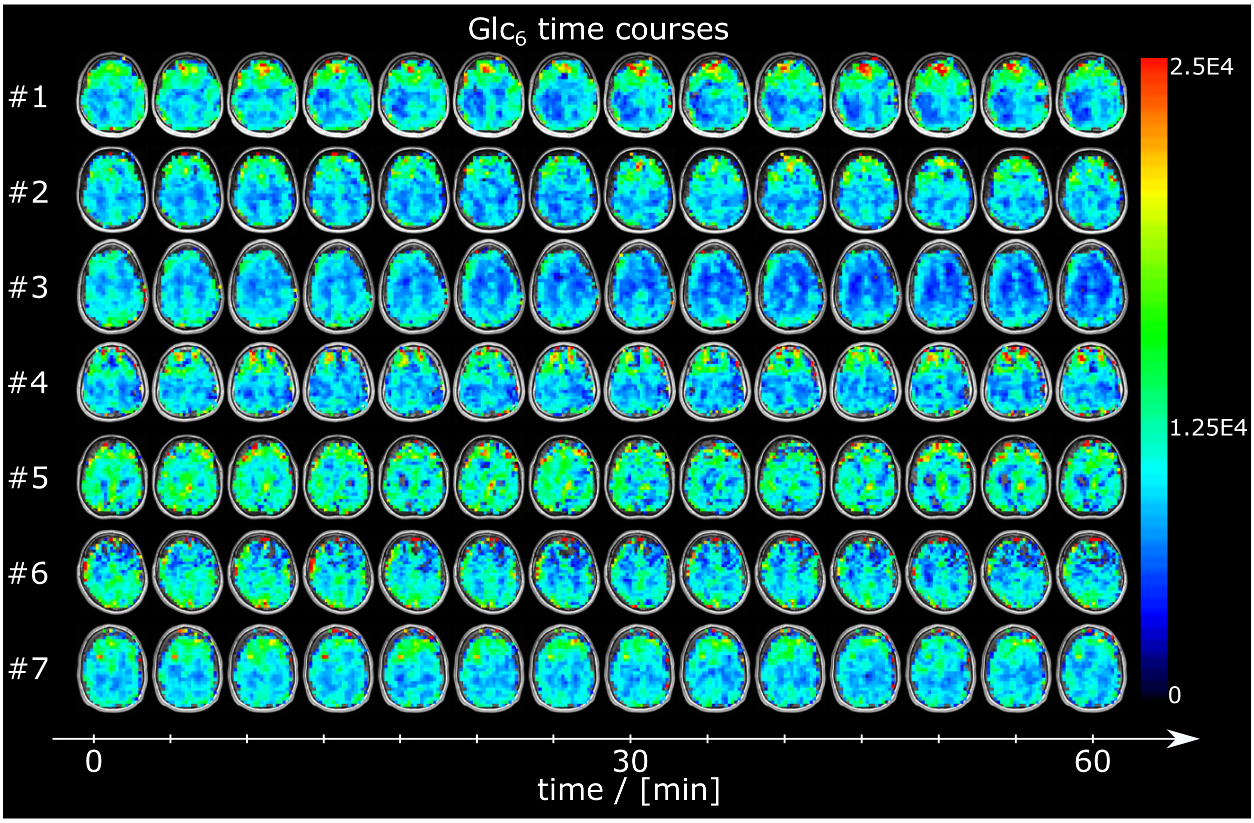


**Supplemental Digital Content Figure 3:** Time courses of axial glucose ($Glc_6$) maps from all subjects over the entire measurement visualizing the image intensity decrease over time due to deuterium labeling. No quality criteria threshold was applied for $Glc_6$ data.
